# Sewer Transport Conditions and Their Role in the Decay of Endogenous SARS-CoV-2 and Pepper Mild Mottle Virus from Source to Collection

**DOI:** 10.1101/2024.10.01.24314490

**Authors:** Élisabeth Mercier, Patrick M. D’Aoust, Walaa Eid, Nada Hegazy, Pervez Kabir, Shen Wan, Lakshmi Pisharody, Elizabeth Renouf, Sean Stephenson, Tyson E. Graber, Alex E. MacKenzie, Robert Delatolla

## Abstract

This study presents a comprehensive analysis of the decay patterns of endogenous SARS-CoV-2 and Pepper mild mottle virus (PMMoV) within wastewaters spiked with stool from infected patients expressing COVID-19 symptoms, and hence explores the decay of endogenous SARS-CoV-2 and PMMoV targets in wastewaters from source to collection of the sample. Stool samples from infected patients were used as endogenous viral material to more accurately mirror real-world decay processes compared to more traditionally used lab-propagated spike-ins. As such, this study includes data on early decay stages of endogenous viral targets in wastewaters that are typically overlooked when performing decay studies on wastewaters harvested from wastewater treatment plants that contain already-degraded endogenous material. The two distinct sewer transport conditions of dynamic suspended sewer transport and bed and near-bed sewer transport were simulated in this study at temperatures of 4°C, 12°C and 20°C to elucidate decay under these two dominant transport conditions within wastewater infrastructure. The dynamic suspended sewer transport was simulated over 35 hours, representing typical flow conditions, whereas bed and near-bed transport extended to 60 days to reflect the prolonged settling of solids in sewer systems during reduced flow periods. In dynamic suspended sewer transport, no decay was observed for SARS-CoV-2, PMMoV, or total RNA over the 35-hour period, and temperature ranging from 4°C to 20°C had no noticeable effect. Conversely, experiments simulating bed and near-bed transport conditions revealed significant decreases in SARS-CoV-2 and total RNA concentrations by day 2, and PMMoV concentrations by day 3. Only PMMoV exhibited a clear trend of increasing decay constant with higher temperatures, suggesting that while temperature influences decay dynamics, its impact may be less significant than previously assumed, particularly for endogenous RNA that is bound to dissolved organic matter in wastewater. First order decay models were inadequate for accurately fitting decay curves of SARS-CoV-2, PMMoV, and total RNA in bed and near-bed transport conditions. F-tests confirmed the superior fit of the two-phase decay model compared to first order decay models across temperatures of 4°C to 20°C. Finally, and most importantly, total RNA normalization emerged as an appropriate approach for correcting the time decay of SARS-CoV-2 exposed to bed and near-bed transport conditions. These findings highlight the importance of considering decay from the point of entry in the sewers, sewer transport conditions, and normalization strategies when assessing and modelling the impact of viral decay rates in wastewater systems. This study also emphasizes the need for ongoing research into the diverse and multifaceted factors that influence these decay rates, which is crucial for accurate public health monitoring and response strategies.

## 1. Introduction

In late 2019, SARS-CoV-2 virus was first detected in the Hubei province of China (Huang et al., 2020; Xiantian et al., 2020). The pandemic saw a global scientific advancement of wastewater-based surveillance (WBS), the surveillance of wastewater to detect and quantify pathogens in wastewaters and to assess transmission of disease in communities (Brouwer et al., 2018; Diamond et al., 2022; Michael-Kordatou et al., 2020; O’Reilly et al., 2020; Park et al., 2020; Peccia et al., 2020; Xu et al., 2020). Since the advancement of WBS, strong correlations have been reported between measured SARS-CoV-2 viral RNA concentrations in municipal wastewaters and traditional public health metrics used to monitor the pandemic’s progress, such as new daily reported clinical cases, percent positivity of clinical tests, hospitalization admissions and deaths caused by COVID-19 complications (Ahmed et al., 2020a; D’Aoust et al., 2022, 2021a; Hegazy et al., 2022; Keshaviah et al., 2023; Kumar et al., 2020; La Rosa et al., 2020; Randazzo et al., 2020; Thompson et al., 2020). As a result, SARS-CoV-2 WBS has since been applied at scale in over 4,648 sites, in at least 72 countries worldwide (Naughton et al., 2023), to help governmental agencies and public health organizations.

Significant efforts and developments have been made to improve the measurement of SARS-CoV-2 in wastewaters and specifically to increase the sensitivity of assays (Pecson et al., 2021). While several comprehensive reviews and analyses on laboratory methodologies have been performed, resulting in improved and robust testing methodologies, there still exists a lack of fundamental understanding of the impacts of sewershed-induced decay on viral signal measurements. In particular, the effects of short, moderate and long sewer transport, and the presence of industrial waste or chemical disinfectants on measured viral titers of SARS-CoV-2 have not yet been well elucidated, and limited literature currently exists on these topics (Parra-Arroyo et al., 2023). Furthermore, at the time of writing, only a limited number of studies (9) have attempted to elucidate decay kinetics of SARS-CoV-2 viral signal in wastewaters (Ahmed et al., 2020b; Babler et al., 2023; Bivins et al., 2020; de Oliveira et al., 2021; Hart et al., 2023; Hokajärvi et al., 2021; Roldan-Hernandez et al., 2022; Sala-Comorera et al., 2021; Weidhaas et al., 2021; Yang et al., 2022), with the reported decay kinetics of the existing studies investigating SARS-CoV-2 viral decay shown in Table 1. Six out of ten studies employed spiked-in SARS-CoV-2 viral particles as opposed to studying endogenous SARS-CoV-2 already present in wastewater. This distinction is significant as the decay rates between endogenous, fecally shed, SARS-CoV-2 RNA in wastewater samples, and lab-propagated SARS-CoV-2 virions, remain largely unexplored and not well understood (Kantor et al., 2021). It is hypothesized that spiked-in SARS-CoV-2 viral particles may decay faster than their endogenous counterparts, as endogenous SARS-CoV-2 has been observed to bind to dissolved organic matter in wastewater matrices leading to slower degradation and increased persistence in the environment (Chatterjee et al., 2023). To best elucidate the decay kinetics of SARS-CoV-2 viral signal in sewers and wastewaters, an endogenous material spike-in approach using infected stool from patients is likely best to limit confounding factors from inherent differences between endogenous SARS-CoV-2 RNA and lab-propagated SARS-CoV-2 virions that impact the interpretation of results. Further, a spike-in approach using infected stool would enable decay to be quantified from the time that the viral titers enter the wastewater matrix, enabling the true decay profile to be analyzed from source to point of collection.

**Table 1:**
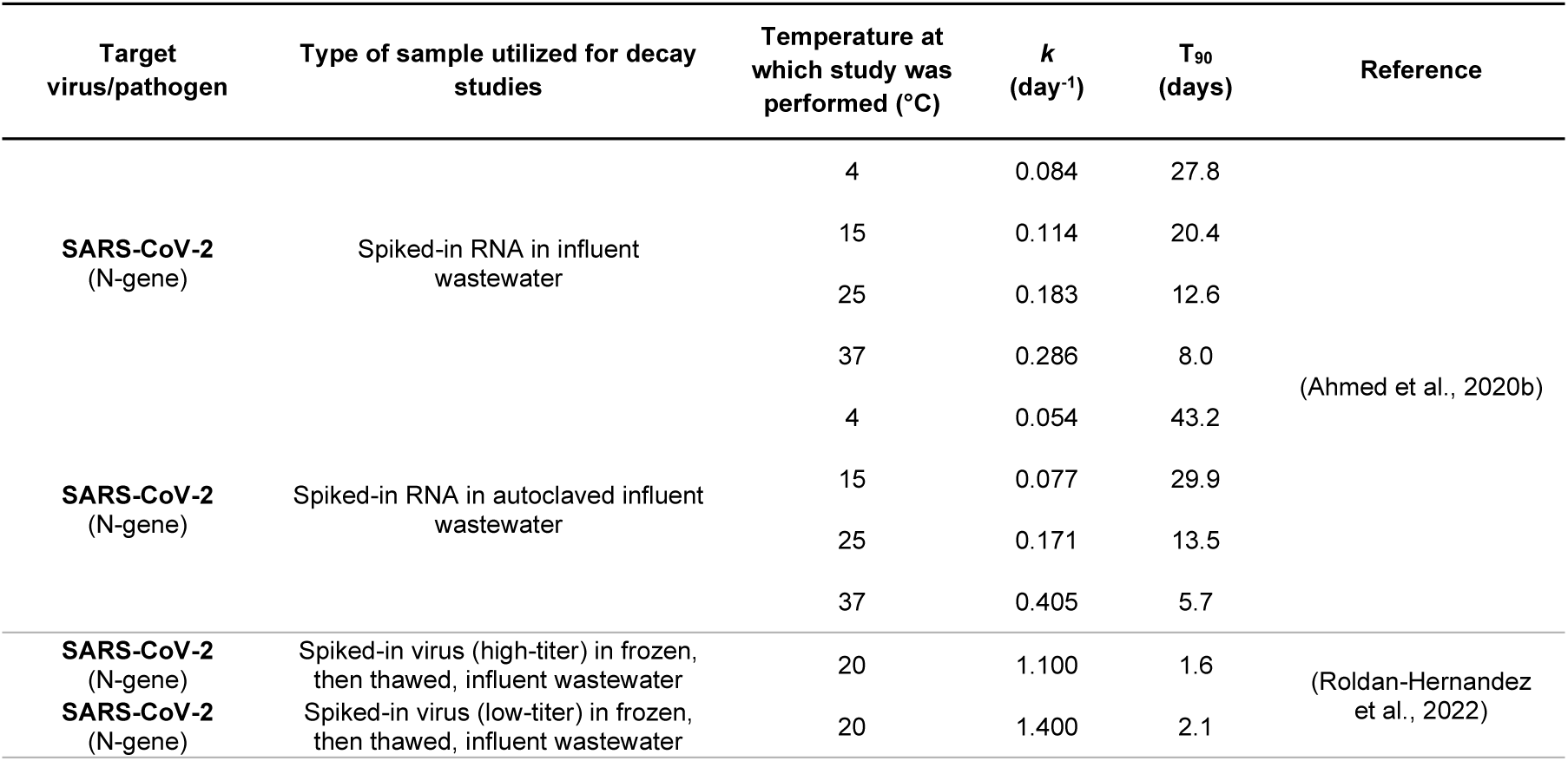
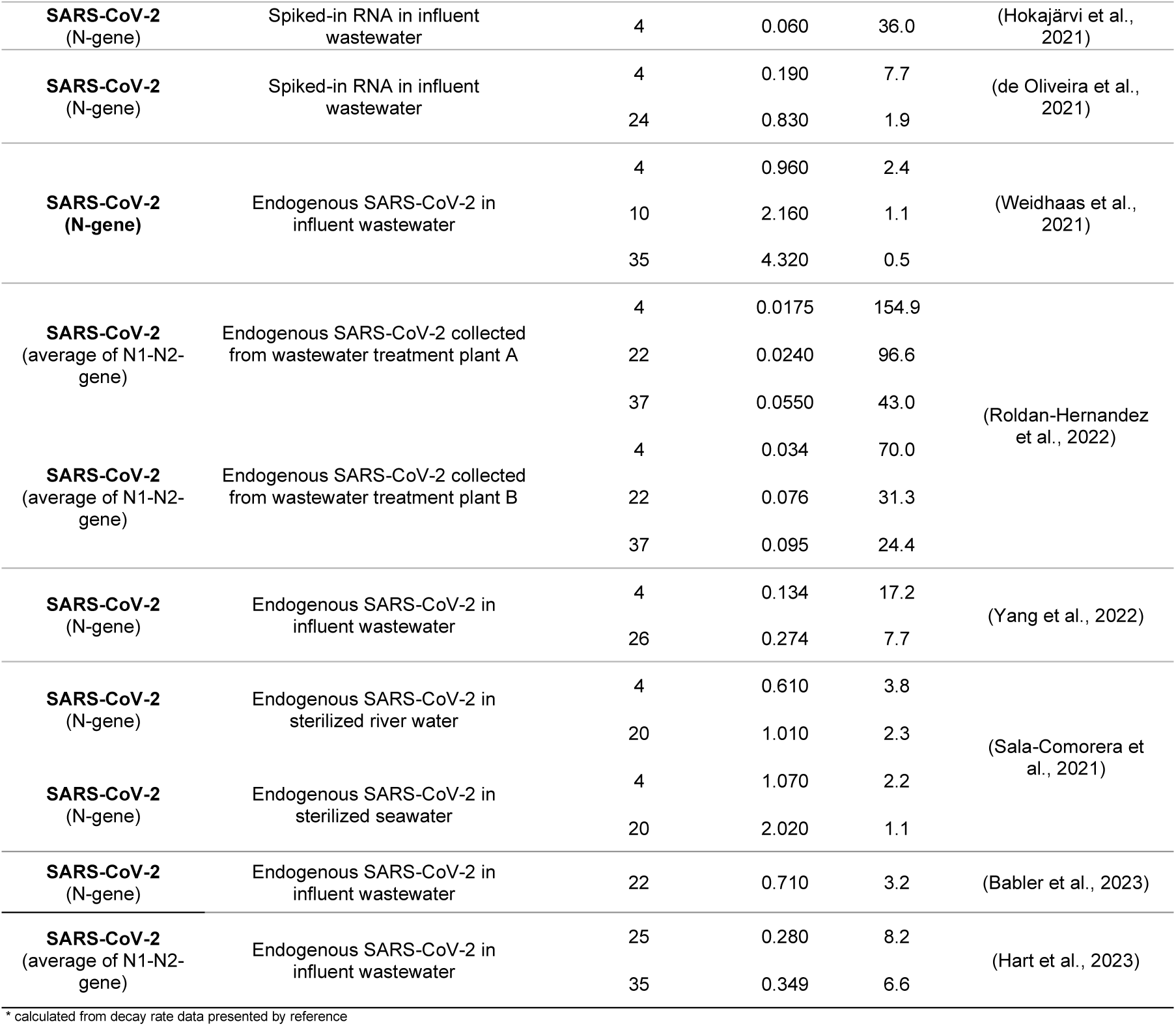
SARS-CoV-2 decay study temperatures, decay kinetics and associated T_90_.

The current range of endogenous decay rates from SARS-CoV-2 positive wastewaters reported in literature is broad, with T_90_ ranging from 2.4 to 154.9 days at 4°C, Table 1 (Roldan-Hernandez et al., 2022; Weidhaas et al., 2021; Yang et al., 2022). While this wide range of values could be somewhat explained by the variating type of starting material, it calls for further investigation into factors such as wastewater matrix composition, titer of the target, and the type of transport the target is subjected to in the sewer system. Additionally, it’s important to note that existing studies using endogenous samples have primarily focused on the decay of endogenous viruses in wastewater samples collected at treatment plants. This approach introduces a significant bias, as these samples have already undergone decay during their residence time in the sewer system, which can extend to more than a day depending on the system, in addition to the holding time prior to the experiment. Consequently, current decay studies may not accurately include the initial decay of the virus titers from the source of entry in the system (such as a toilet flush), leading to a biased understanding of the decay kinetics. Furthermore, existing studies do not incorporate considerations for different flow dynamics existing in the sewersheds, as all studies to date on SARS-CoV-2 viral signal decay in wastewater have been performed in a manner that most closely mimics bed and near-bed sewer transport, which could impact the interpretation of results, and might be limiting WBS applications such as public health reporting, or modelling.

A common fecal biomarker, pepper mild mottle virus (PMMoV), has been used by approximately 30% of SARS-CoV-2 surveillance systems worldwide to normalize reported viral signal to the quantity of fecal matter in the samples (D’Aoust et al., 2021a; Feng et al., 2021; Haramoto et al., 2013; Kitajima et al., 2018; Naughton et al., 2023). As fecal normalization with PMMoV directly impacts reported normalized SARS-CoV-2 viral signal measurements, it is equally important to understand the effects of decay on PMMoV, if any, throughout the sewershed. The reported decay kinetics of the existing studies investigating PMMoV viral decay are shown below in Table 2. Similar to SARS-CoV-2, the current range of decay rates reported for PMMoV is quite broad, ranging from 57.6 to 237 days at 4°C raising the same concern for further investigation.

**Table 2:** PMMoV decay study temperatures, decay kinetics and associated T_90_.

| Target virus/pathogen | Type of sample utilized for decay studies | Temperature at which study was performed (°C) | k (day<sup>-1</sup>) | T<sub>90</sub> (days) | Reference |
| --- | --- | --- | --- | --- | --- |
| **PMMoV (RAP-gene)** | Endogenous PMMoV in wastewater/wetland water | 4 | 0.040 | 57.6* | (Rachmadi, 2016) |
|  |  | 25 | 0.050 | 46.1* |  |
|  |  | 37 | 0.080 | 28.8* |  |
| **PMMoV (RAP-gene)** | Endogenous PMMoV in river water | 20 | 0.228** | 10.2 | (Sala-Comorera et al., 2021) |
| **PMMoV (RAP-gene)** | Endogenous PMMoV in wastewater (WRRF 1) | 4 | 0.010 | 237.4 | (Roldan-Hernandez et al., 2022) |
|  |  | 22 | 0.040 | 57.2 |  |
|  |  | 37 | 0.045 | 51.7 |  |
| **PMMoV (RAP-gene)** | Endogenous PMMoV in wastewater (WRRF 2) | 4 | 0.059 | 39.0 |  |
|  |  | 22 | 0.077 | 30.1 |  |
|  |  | 37 | 0.091 | 25.3 |  |
| **PMMoV (RAP-gene)** | Endogenous PMMoV in influent wastewater | 22 | 0.453 | 5.1* | (Babler et al., 2023) |

WRRF = Water Resource Recovery Facility, \* calculated from decay rate data presented by reference, \*\* calculated from data presented by Sala-Comorera (2021)

Other normalizers used in WBS studies include various RNA targets. RNA viruses have commonly been used as spiked-in controls for assessing extraction efficiency in WBS studies (Torii et al., 2022). Additionally, RNA extracts have been employed as spike-in inhibition controls during the qPCR step of sample processing (Ahmed et al., 2020c). Given its established role as a control in extraction and qPCR processes, RNA’s utility in wastewater surveillance is well recognized. This role, coupled with the established prevalence of human-associated viruses and bacteria-infecting viruses contributing to the urban virome in sewage, supports total RNA concentration as a robust candidate for normalizing against decay and temperature effects (Guajardo-Leiva et al., 2020; Nieuwenhuijse et al., 2020; Tisza et al., 2023). The presence of diverse RNA families in sewage indicates that this normalization technique could be effective for multiple endogenous sewage viral targets, particularly if they exist in sewage primarily as components of broken virions, rather than as intact virions with various structural properties.

Numerous concurrent biological processes occur in wastewater sewers, these processed may include both aerobic and anoxic/anaerobic biochemical reactions, depending on flow conditions. Two of the most common flow conditions associated with constituent transport in sewers are: i) dynamic suspended sewer transport, and ii) bed and near-bed transport. Dynamic suspended sewer transport conditions occur during conventional, dynamic, design-flow conditions within the sewers and are characterized as quasi-fully mixed flows with suspended solids transport. This transport condition occurs during conventional weather and during conventional downstream operation of wastewater treatment facilities. It is distinguished from other modes of sewer transport conditions by the lack of pronounced solids deposition and a high degree of oxygen entrainment into the liquid sewage matrix, creating semi-aerobic to aerobic conditions, along with significant particle movement (Bertrand-Krajewski et al., 2010; Qteishat et al., 2011). Wastewaters that are subjected to dynamic suspended sewer transport remain within the sewershed for the design residence time of the sewershed, with conventional sewersheds rarely exceeding past a few days of residence time. Bed and near-bed transport conditions are characterized by an accumulation of deposited wastewater solids within the sewershed and is caused by relatively low flow velocities of the wastewaters, with the flow velocities falling below those required for continued solids suspension (Bertrand-Krajewski et al., 2010). Bed and near-bed transport conditions are cyclical in nature, and most often occur in regions of the sewershed where flow velocities are unable to be constantly maintained in a manner ensuring suspended sewer transport conditions (Ashley and Crabtree, 1992; CRABTREE, 1989; Lange and Wichern, 2013). Notably, this phenomenon occurs in seasonal climates where yearly cycles of low and high rainfall seasons are present. Similarly, in northern and cold climate countries, precipitation accumulates as snow and ice during colder months, representing periods of low flow and sedimentation for months, until the warmer months bring increased flow, thus reinstating higher velocity conditions in the sewers. Finally, within some municipalities, bed and near-bed transport conditions may be induced when the sewers are used to store wastewaters during maintenance activities at the downstream wastewater treatment facility.

A limited understanding of the persistence and degradation of viral material in wastewaters remains due to a lack of decay studies that use infected stool spiked into wastewaters and hence studies that investigate of decay rates from the time of individual contributions to community-level sampling. This considerable limitation in our current knowledge coupled with the significant variation in reported decay rates of spiked-in SARS-CoV-2 viral particles studies and endogenous SARS-CoV-2 already present in wastewater studies warrants further investigation of the persistence and degradation of endogenous SARS-CoV-2 viral material in wastewaters. Our study addresses these gaps by examining the decay rates of SARS-CoV-2 and PMMoV from the point of entry into the sewer system by using infected stool spiked into wastewater and studying persistence and degradation of the viral material under the two common sewer transport conditions of dynamic suspended transport and bed and near-bed transport. This approach leverages the advantages of spike-in studies for evaluating decay from time zero while benefiting from the use of endogenous infected material while also simulating realistic environments within wastewaters through the replication of common transport conditions. This method hence more accurately mirrors the natural conditions the virus encounters upon entering the sewer system. We hypothesize that the decay dynamics of pathogens within sewer systems are significantly influenced by both the flow conditions and the hydraulic retention time, the latter being intrinsically linked to urban planning and population density, which consequently impact the outcomes of WBS across different municipalities. These factors are critical for interpreting viral signal variations across regions and can facilitate a more global understanding of pathogen prevalence and movement. By studying the decay rates from the initial stages and within more accurate sewer environments, our research aims to develop a more precise and comprehensive understanding of viral persistence and decay dynamics in wastewater systems, thus contributing to the enhancement of wastewater-based epidemiological models.

## 2. Materials and Methods

### 2.1. Stool and wastewater samples

SARS-CoV-2 positive stool samples containing endogenous SARS-CoV-2 viral material were spiked into wastewater containing endogenous SARS-CoV-2 to perform the decay experiments, thus simulating the contribution from source (flushed toilet) into wastewater. To determine the decay rate starting at the time of entry in the sewershed, 5 anonymous stool samples were obtained from consenting SARS-CoV-2 infected adult patients, combined into a paste, and used to spike the wastewater. The only information obtained from the contributing patients was a confirmed positive COVID-19 test result, with no other personal medical records obtained. Hence the collection of stool samples for this study was exempt from research ethics board review. Stool samples were transported on ice and stored at 4°C in the laboratory. Upon arrival, five biological replicates of stools were immediately extracted and screened for SARS-CoV-2 and PMMoV using RT-qPCR, as described below, to quantify the level of endogenous viral signals in the spiking material.

An 18.9 L grab sample of post-grit influent wastewater was collected for this study from the City of Ottawa’s Robert O. Pickard Environmental Center, the city’s only wastewater treatment plant, which receives and treats the wastewater of approximately 91% of the residents in Ottawa (Supplemental Table S1). The collected wastewater sample was transported to the laboratory on ice and preserved at 4°C until the start of the decay experiments and was not pasteurized or otherwise altered in any way. Upon arrival, five biological replicates of the wastewater were immediately extracted and screened for SARS-CoV-2 and PMMoV using RT-qPCR, as described below, to quantify the level of endogenous viral signals. Decay experiments began within 12 hours of the collection of the wastewater. A grab sample was selected to prioritize the freshness of the sample over representativeness, as the study did not require monitoring of diurnal variations, and using a composite sample would have introduced an additional 24 hours of potential decay before analysis.

### 2.2. Temperature-controlled experimental chambers

To conduct decay experiments at various, constant temperatures, three small refrigerators were temperature-controlled to specific experimental temperatures using an external Inkbird ITC-308 Digital Temperature Controller to act as temperature-controlled experimental chambers. The three small refrigerators were maintained at the temperatures of 19.6°C ±1.0°C, 12.5°C ±1.1°C and 4.9°C ± 0.6°C, throughout all decay experiments, respectively.

### 2.3. SARS-CoV-2, PMMoV and total RNA decay experiments simulating dynamic suspended transport conditions at three distinct temperatures

**Figure 1:**
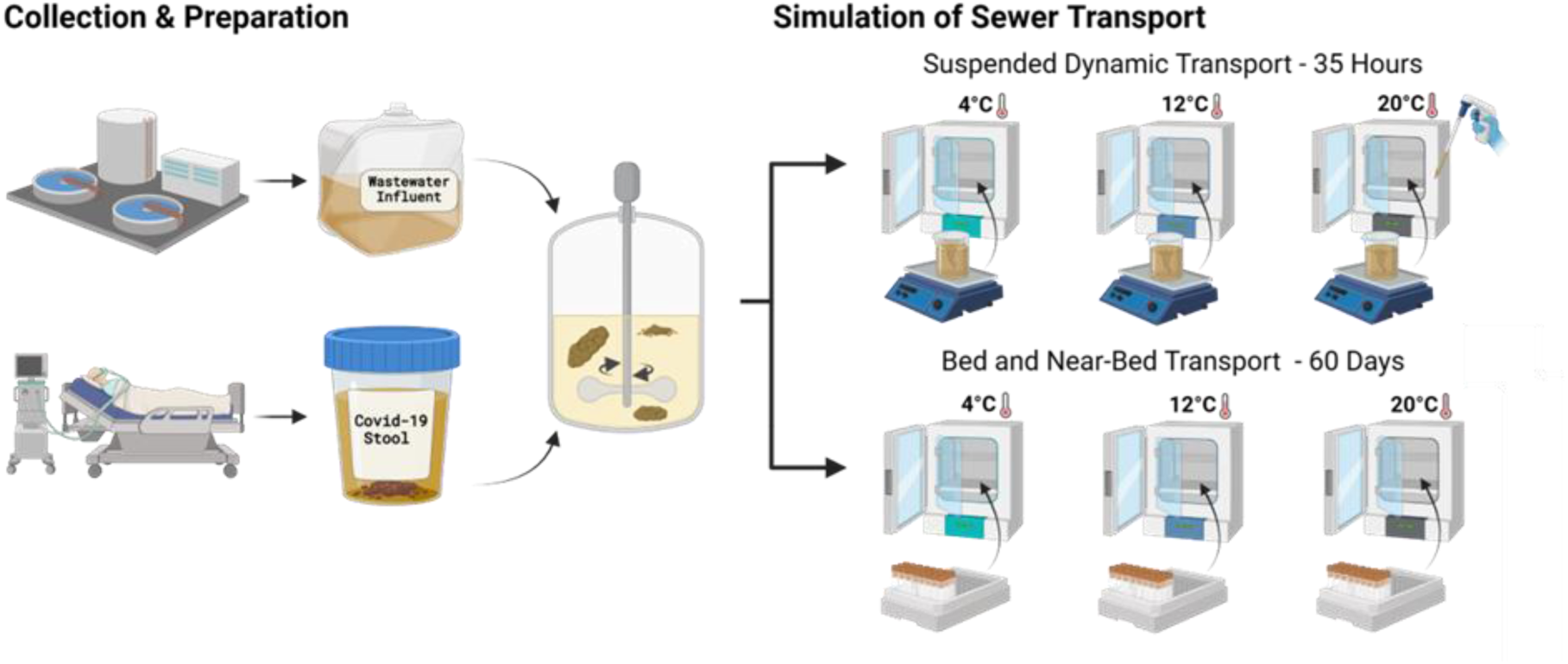
Experiment set up simulating suspended and bed and near-bed transport conditions in sewersheds.

A series of dynamic suspended transport SARS-CoV-2, PMMoV and total RNA decay experiments were performed to simulate sewer transport of viral material within small, medium and large subsections of sewersheds. Hence, short, moderate and long sewer hydraulic retention time times of 0 hours, 2.5 hours, 6.0 hours, 15.0 hours, 24.0 hours, and 35.0 hours were used in this study to reflect the varying distances wastewater travels, based on the distance from its source, such as households, to the treatment plant. 17.61 grams of stool were dissolved into 2.40 liters of post-grit wastewater, achieving a concentration of approximately 7.34 g/L, and the mixture was carefully agitated at 4°C until fully homogenized. The stool-wastewater mix was then separated into three (3) individual reactors containing magnetic stir bars and placed within temperature-controlled experimental chambers set at target temperature of 4° C, 12° C and 20°C, respectively. The lowest temperature, 4° C, was specifically selected to reflect the conditions in sewers of northern climate countries during colder periods, while the 12° C and 20°C settings are representative of a wider range of temperatures commonly encountered in sewer systems across various regions globally (Hart and Halden, 2020; Vialkova et al., 2020; Wilson and Worrall, 2021). Magnetic stir-plates were installed inside the temperature-controlled experimental chambers and the 3 individual vessels containing the stool-wastewater mix were placed on top of the stir-plates, where the magnetic stir plates were used to simulate the well-mixed, dynamic movement and suspension conditions of wastewater-associated solids and fecal material during suspended transport conditions. Five representatives 40 mL samples of well-homogenized stool-wastewater mixtures were harvested from each reactor vessel at 0, 2.5, 6.0, 15.0, 24.0, and 35.0 hours. Each time point had five biological replicates, and at the end of the experiment, half of the initial volume remained in the reactor to ensure representative conditions.SARS-CoV-2 RNA, PMMoV RNA and total RNA were extracted immediately after sampling. Extracted RNA was never frozen and instead kept at 4° C until RT-qPCR analysis was performed within 48 hours of extraction, a timeframe during which literature has confirmed RNA stability(Robinson et al., 2021; Torabi et al., 2023). Control experiments with wastewater samples that did not contain spiked stool were conducted under identical conditions and are shown in Supplemental Figure S1. This approach was to verify that the spiked material from infected patients, although endogenously similar to the material found in typical wastewater samples, behaved comparably to samples collected directly from the treatment plant only 12 hours earlier.

### 2.4. SARS-CoV-2, PMMoV and total RNA decay experiments simulating bed and near-bed transport conditions at three distinct temperatures

A series of SARS-CoV-2, PMMoV and total RNA decay experiments were designed to simulate bed and near-bed transport conditions in conventional sewersheds, including sedimentation time representative of wintertime flow conditions in northern and cold climate countries, where precipitation and groundwater infiltration effects on the sewershed are significantly reduced. 48 grams of stool were dissolved in 7 liters of post-grit wastewater, achieving a concentration of approximately 6.86 g/L, and were carefully agitated at 4°C until the stool was fully mixed into the wastewater. This slightly lower stool concentration was due to logistical constraints in measuring and dissolving the stool such as residues adhering to the plastic weighing boats. The stool-wastewater mix was then transferred to forty-five (45) individual 50 mL conical centrifuge tubes. Three individual temperature-controlled experimental chambers were set to 4° C, 12° C and 20 °C, respectively, and were used to house the 50 mL conical centrifuge tubes, which were not agitated, simulating quiescent sewershed conditions and sedentary bed and near-bed transport of wastewater-associated solids and fecal material. Five individual 50 mL conical centrifuge tubes containing stool-wastewater mixtures were harvested at periodic intervals between 0 and 60 days from each temperature-controlled experimental chamber. Extended sampling for simulating bed and near-bed transport conditions was necessary due to these conditions persisting for months. Indeed, sediment layers can remain for months in the sewer system until eroded by environmental factors (Lange and Wichern, 2013). In colder climates like Ottawa, precipitation solidifies as snow and ice, reducing flow and erosion until warmer months, when snowmelt and rainfall initiate erosion. Samples were extracted immediately after sampling, extracted RNA was never frozen and instead maintained at 4° C until, with RT-qPCR analysis being performed within 48 hours of collection of the sample. Control experiments with wastewater samples that did not contain spiked stool were conducted under identical conditions and are shown in Supplemental Figure S2. This approach was to verify that the spiked material from infected patients, although endogenously similar to the material found in typical wastewater samples, behaved comparably to samples collected directly from the treatment plant only 12 hours earlier.

### 2.5. Sample concentration and nucleic acid extraction

Representative 40 mL samples of well-homogenized stool-wastewater mixtures were collected from the suspended and bed and near-bed transport temperature-controlled experimental chambers and were immediately processed. Samples were concentrated and SARS-CoV-2, PMMoV, and total RNA were extracted as described by D’Aoust et al. (2021b). Briefly, samples were centrifuged at 12,000 x g for 45 minutes, and the supernatant was discarded. Samples were then centrifuged once again at 12,000 x g for an additional 5 minutes, and the resulting supernatant was again discarded. RNA was extracted from the resulting pellet using Qiagen’s RNeasy PowerMicrobiome Kit (Qiagen, Germantown, MD), with the following modifications to the manufacturer’s procedures: i) 250 mg of the resulting solids pellet was extracted instead of a 200 µL liquid sample, and ii) the optional phenol-chloroform solution was substituted by Trizol LS reagent (ThermoFisher, ON, Canada). Samples were then eluted in 100 µL of RNAse/DNAse-free water.

### 2.6. RT-qPCR SARS-CoV-2 and PMMoV analyses

The SARS-CoV-2 viral signal was quantified using a singleplex one-step, RT-qPCR targeting the N1 and N2 genomic regions. The PMMoV viral signal was also measured using a singleplex one-step with RT-qPCR targeting a region in the replication-associated protein portion of the genome (Haramoto et al., 2013). Each PCR reaction was composed of 1.5 µL of RNA template, forward and reverse primers (final concentration of 500 nM each), probe (final concentration of 250 nM) 2.5 µL of 4x TaqMan^®^ Fast Virus 1-step Mastermix (ThermoFisher, ON, Canada), in a total reaction volume of 10 µL. All primer sequences used in this study are shown in Supplemental Table S2. All samples were run in technical triplicates with non-template controls and 5-point standard curves prepared with the Exact Diagnostic (EDX) COV019 SARS-CoV-2 RNA standard (Exact Diagnostics, TX, USA). PCR cycling conditions were identical as those described previously (D’Aoust et al., 2021b). The assay’s limit of detection (ALOD) and quantification (ALOQ) for SARS-CoV-2’s N1 region were of approximately 2 copies/reaction and 3.2 copies for reaction, respectively. For the N2 region, they were of approximately 2 copies/reaction and 8.1 copies/reaction, respectively. All cycling conditions used in this study are shown in Supplemental Table S3.

### 2.7. Total RNA analysis

The total RNA concentration of each sample was measured using an Agilent 2100 Bioanalyzer. 2 µL of extracted RNA elution was loaded on an RNA 6000 Pico Chip. Further data analysis and concentration determinations were performed using Agilent’s 2100 Expert software (v. B.02.10.SI764).

### 2.8. Sanger sequencing of amplicons

Sanger sequencing was conducted on time point day 21, day 45 and day 60, to ensure that the analyses did not produce false positives. The specificity of the amplicons resulting from RT-qPCR analyses generated for the various targets of this study was evaluated via Sanger sequencing. First, a touchdown PCR (TD-PCR) was performed using Q5^®^ High-Fidelity DNA Polymerase with 1 µL of RT-qPCR amplicons as the starting template. The initial touchdown was performed as follows: [98 °C (30 seconds) + 64 °C → 55 ^°^C, drop of 1 °C/cycle, + 72 °C (30 seconds)] x 10 cycles. Amplification was then performed as follows: [98 °C (30 seconds) + 64 °C → 55 ^°^C, drop of 0.4 °C/cycle, + 72 °C (30 seconds)] x 25 cycles. The TD-PCR products were then run on a 3% agarose gel at 100V to separate the amplicons. The amplicon band observed at the appropriate location was then cut and gel extracted using Monarch^®^ DNA Gel Extraction Kit (New England Biolabs, MA, USA) as per the manufacturer’s instructions. After obtaining purified DNA, a novel primer extension strategy for one-step PCR amplification was performed using the Q5^®^ High-Fidelity DNA Polymerase. In brief, 1 ng of DNA was used to extend the N1 and N2 amplicon using the oligo adapters (Supplemental Table S2) with partial complementarity to the targeted amplicons. An extension PCR amplification was then performed as follows: 98 °C (30 seconds) + [98 °C (10 seconds) + 50 °C (30 seconds) + 72 °C (30 seconds) x 30 cycles, + 72 °C (2 minutes). The extension-PCR amplified product is then purified using QIAquick^®^ PCR Purification Kit (Qiagen, MD, USA) following the manufacturer’s instructions. The final amplicon product was then sequenced by Sanger Sequencing at the Ottawa Hospital’s Research Institute (OHRI) StemCore Sequencing Facility using an ABI Prism 3730 DNA Sequencer (Applied Biosystems, MA, USA). The sequences were compiled and edited using BioEdit (ver. 7.2)(Hall, 1999) and sequence alignment was done by Clustal Omega (Madeira et al., 2022). Each PCR reaction had a total volume of 25 µL and was composed of the amplicons’ regular reverse primers (500 nM), A target-specific amplicon-seq-Stuffer forward primer (50 nM), Stuffer-1 forward primer (50 nM), Stuffer-2 forward primer (500 nM), dNTP (200 µM), and 1X of the 5X Q5 reaction buffer and Q5 high fidelity DNA polymerase (0.02U/µL). All primer sequences used in this study are shown in Supplemental Table S2.

### 2.9. Volume, PMMoV and total RNA normalization

Measurements of SARS-CoV-2 in this study were normalized using volume, PMMoV, and total RNA to investigate potential degradation mitigation techniques. Volume normalization was employed instead of flow normalization to accurately reflect the conditions of this controlled experiment, where sampling was conducted from a batch reactor rather than a continuous flow system. As such, volume normalization serves as an extrapolation of flow normalization typically used in wastewater treatment plants with variable daily flow rates. Additionally, since SARS-CoV-2 and PMMoV are known to be prevalent in the solids fraction (D’Aoust et al., 2021a), volume normalization in this study effectively simulates the real-world scenario where, despite consistent sampling volumes, the amount of solids and consequently, the viral signal can vary, even when flow normalization is applied.

### 2.10. Statistical analyses

#### First order decay rate

The first order decay rate model was applied to the SARS-CoV-2, PMMoV and total RNA targets under dynamic suspended transport conditions and bed and near-bed transports at temperatures of 4° C, 12° C and 20 °C. The first order decay rate constant (k) was calculated as shown in equation 1 (Chick, 1908). Here, the term [*A*]_*t*_ represents the concentration of the target at time t, while [*A*]_0_ is the initial concentration at time zero. The rate was obtained by calculating the slope of the natural logarithm (Ln) of the concentration of the target’s (A) signal versus time (t). The slope was calculated using GraphPad Prism (version 9.3.1).

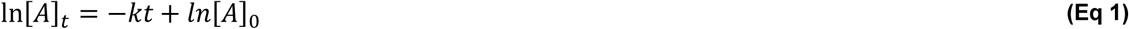

#### Significance of first order decay rate constant

To assess whether the SARS-CoV-2, PMMoV and total RNA targets exhibited decay, a two-tailed t-test with a significance level (α) of 0.05, was used to determine the significance of the first order decay constant under dynamic suspended transport conditions and bed and near-bed transport conditions at temperatures of 4° C, 12° C, and 20 °C, with the null hypothesis that the decay rate is zero. These tests were performed on k values obtained from the first order decay model for all targets at each temperature to determine if each k was significantly different from zero.

#### Comparison of first order decay rate constant

Differences in decay rate constants between targets at various temperatures were assessed using a two-tailed t-test with a significance level (α) of 0.05, with the null hypothesis that the decay rates of the targets were not significantly different. Specifically, three separate t-tests were conducted for each comparison, always comparing two groups at a time: SARS-CoV-2 vs. PMMoV, SARS-CoV-2 vs. total RNA, and PMMoV vs. total RNA. Additionally, three independent t-tests were performed to compare the normalized signals (volume-normalized vs. PMMoV-normalized, volume-normalized vs. total RNA-normalized, and PMMoV-normalized vs. total RNA-normalized) at temperatures of 4°C, 12°C, and 20°C. Finally, three independent t-tests were conducted to compare decay rates between different temperature conditions (4°C vs. 12°C, 4°C vs. 20°C, and 12°C vs. 20°C). All t-tests were performed using GraphPad Prism (version 9.3.1).

#### First order decay rate time needed to achieve 90% reduction (T_90_)

T_90_, the time required for 90% of the starting target to decay, was calculated for the first order decay model as shown in equation 2. All T_90_ values were utilized as a comparative measure for analyzing the SARS-CoV-2, PMMoV and total RNA decay rates at temperatures of 4° C, 12° C and 20 °C and for comparisons with other studies.

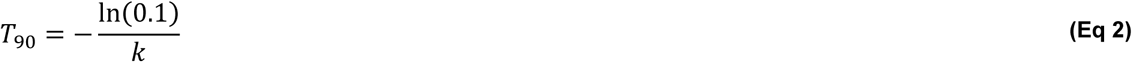

#### Model fit

To assess model fit, the coefficient of determination (R^2^) was calculated for both the first order and two-phase models for all targets and temperatures of the study using GraphPad Prism (version 9.3.1). The R^2^ was used to calculate the proportion of the variation in the dependent variable predictable from the variation in the independent variable (Gage, 1988).

#### Two-phase decay rate

The two-phase decay rate model was applied to the bed and near-bed transport condition data sets to achieve better model fit compared to the first phase decay model. The two-phase decay constants (k_fast_ and k_slow_) were calculated using a two-phase decay model as defined in equations 3 to 3.2 using GraphPad Prism (version 9.3.1). This composite exponential decay model defines the overall decay rate as the sum of a simultaneous fast and a slow exponential decay as shown in equation 3. The “Percent Fast” parameter defines the proportion of the initial concentration [*A*]_0_ subjected to the fast decay process, characterized by the rate constant k_fast_. Simultaneously, the remaining fraction (100 - Percent Fast), undergoes decays at a slower rate k_slow_. The equations 3.1 and 3.2 define the initial concentrations for the fast and slow decay phases, respectively, which are then incorporated into the two-phase decay model. GraphPad Prism offers the option to include a non-zero plateau, representing the terminal concentration at which the decay stabilizes. Based on the nature of our study and the expectation that the concentration diminishes entirely over time, we set the plateau to zero.

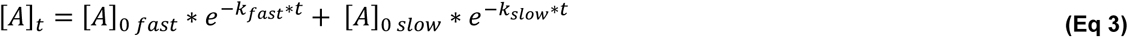

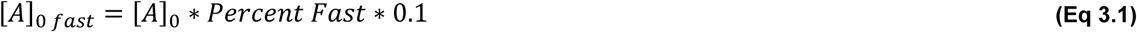

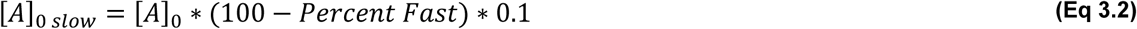

#### Two-phase decay rate time needed to achieve 90% reduction (T_90_)

T_90_, the time required for 90% of the starting target to decay, was calculated for the two-phase decay model as shown in equation 4. T_90_ was calculated for two-phase decay model by solving equation 4 for t using a bisection numerical method. This calculation was performed using the “uniroot” function from the “rootSolve” library in R programming language. All T_90_ values were utilized as a comparative measure for analyzing the decay rates of SARS-CoV-2, PMMoV and total RNA decay rates at temperatures of 4° C, 12° C and 20 °C.

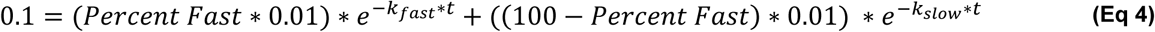

## 3. Results and Discussion

### 3.1. Decay of SARS-CoV-2, PMMoV and total RNA under conditions simulating toilet flushed stool and dynamic sewer suspended transport

The dynamic suspended transport experiments, conducted over a 35-hour period, were designed to mimic the rapid transit conditions typically experienced in sewer systems under normal flow conditions, simulating the movement of viral material from the time of a toilet flush using spiked-in infected stool material. Measurements of SARS-CoV-2, PMMoV and total RNA (Figure 2A to 2C), as well as the volume-normalized, PMMoV-normalized and total RNA-normalized SARS-CoV-2 signal (Figure 2D to 2F), are presented over a simulated sewer transport time of 35 hours. The volume-normalized results are calculated by volume normalizing the study data with the reactor volumes, which is analogous to flow-normalized data collected from a full-scale, continuous flow operating system. No significant decay was observed in the SARS-CoV-2, PMMoV and total RNA measurements within 35-hour period. Consequently, there were also no changes observed in the volume-normalized, PMMoV-normalized and total RNA-normalized SARS-CoV-2 signals. These observations, particularly for SARS-CoV-2 and PMMoV, are consistent with previous studies. While the previous studies did not directly replicate dynamic flow conditions, they assessed decay at temperatures between 4°C and 20°C using spiked-in viral materials and endogenous viral material already present in wastewater. Sala-Comorera et al. (2021) employed spiked-in viral material in river water and seawater at 4°C and 20°C, observing no decay in river water at either temperature and no decay in seawater at 4°C, with significant decay occurring only in seawater at 20°C. Similarly, Roldan-Hernandez et al. (2022) used endogenous material from wastewater that had already been in the sewer system for 17 hours and observed little decay during the first 24 hours at temperatures ranging from 4°C to 22°C. Interestingly, there are also contracting findings in the current literature, with studies by Weidhaas et al. (2021) reporting significant decay of endogenous material during studies on sample storage at 4, 10 and 35 C, hence indicating that there may exist other factors influencing decay. As such, this study shows that endogenous SARS-CoV-2 viral material, PMMoV viral material and total RNA released from stool do not significantly decay under suspended sewer transport conditions during conventional sewer travel times from the point of entry in sewersheds to the sampling point at temperatures between 4°C and 20°C.

**Figure 2:**
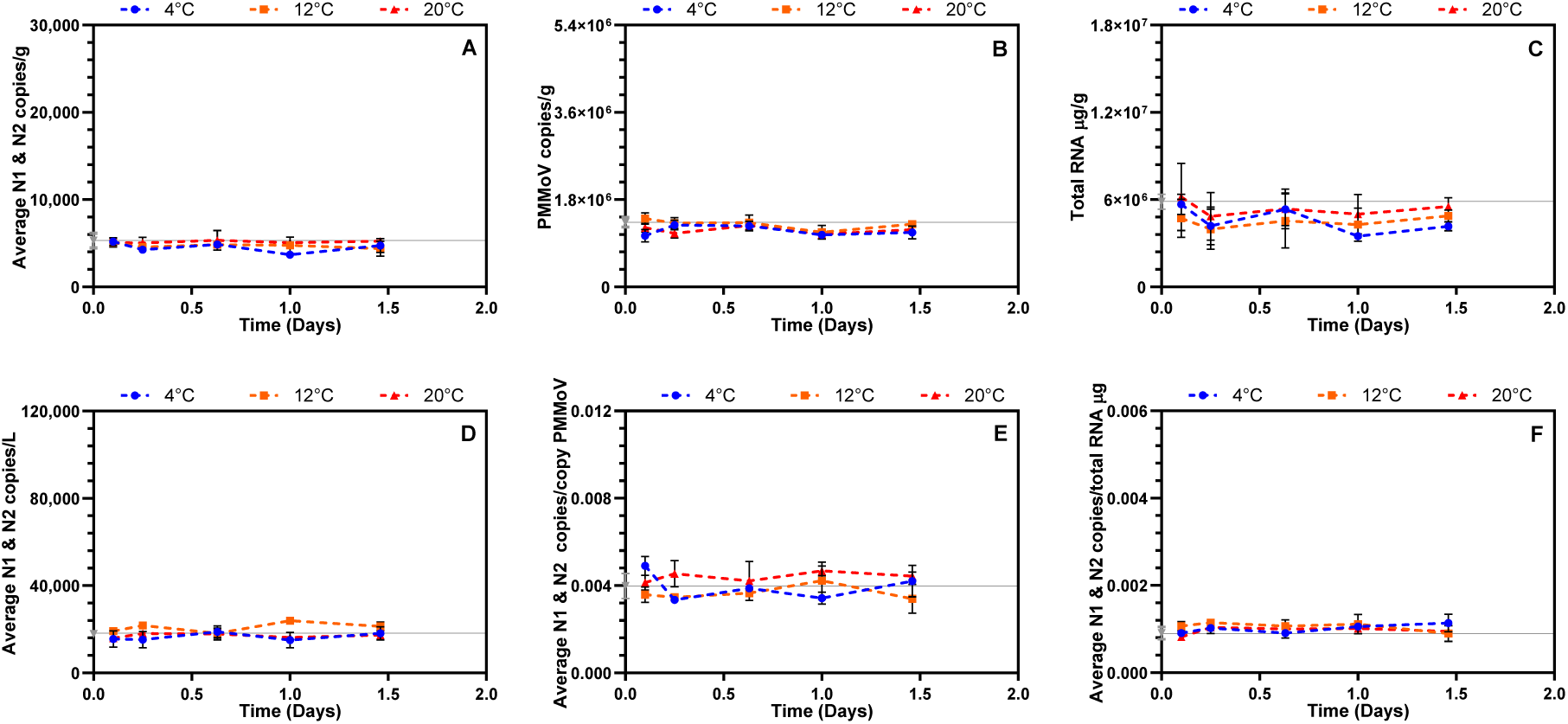
Observed measurements across 35 hours under conditions simulating conventional sewer flow conditions of **A** SARS-CoV-2; **B** PMMoV; **C** total RNA signal. Observed measurements of SARS-CoV-2 across 35 hours under conventional sewer flow setting normalized by **D** volume; **E** PMMoV; **F** total RNA. Starting signal levels are represented by the gray line. Mean and standard deviation are displayed for three temperatures, 4°C, 12°C and 20°C. Where the standard deviation is too small, the error bars are not displayed. Each measurement was performed using 6 technical triplicates from three biological replicates (n=5).

To confirm that no statistically significant differences existed between the viral signal trends over the 35-hour simulated transport period, we conducted a series of independent student’s t-tests. For each targets (3) and normalization methods (3), we assessed the effect of temperature using three separate tests: 4°C vs. 12°C, 4°C vs. 20°C, and 12°C vs. 20°C, resulting in a total of 18 tests. Additionally, we performed t-tests to compare the different normalization methods (volume-normalized vs. PMMoV-normalized, volume-normalized vs. total RNA-normalized, and PMMoV-normalized vs. total RNA-normalized). The resulting p-values were all above the significance threshold (α = 0.05), indicating that neither temperature nor normalization method had a significant effect on persistence or degradation, which was expected given the nonsignificant decay trends observed across all measured targets. These findings are consistent with to those reported by a recent endogenous PMMoV and SARS-CoV-2 decay study by Roldan-Hernandez et al. (2022) that found limited decay when subjected to temperature of 4°C and 22°C for a 10-day period. In contrast, several other studies that use spiked virus with testing conditions that more closely simulate bed and near-bed transport that report an increasing decay rate constant with increasing temperature, especially with temperatures above 25°C (Ahmed et al., 2020b; de Oliveira et al., 2021; Roldan-Hernandez et al., 2022; Weidhaas et al., 2021; Yang et al., 2022). Our study hence addresses gaps in current knowledge and contradictory findings in the literature by demonstrating that SARS-CoV-2, PMMoV and total RNA do not significantly decay under suspended sewer transport conditions after the flush event at temperatures between 4°C and 20°C.

### 3.2. Decay of SARS-CoV-2, PMMoV and total RNA under conditions simulating toilet flushed stool and bed and near-bed sewer transport

The bed and near-bed transport experiments extended up to 60 days to represent the longer retention times of sedimented solids associated with lower flow conditions in the sewer system, particularly during colder months when flow rates decrease. As with the dynamic transport experiments, spiked-in infected stool material was used to simulate the transport of viral material from the time of a toilet flush. The concentrations of SARS-CoV-2, PMMoV and total RNA, Figure 3A to 3, as well as the volume-normalized, PMMoV-normalized and total RNA-normalized SARS-CoV-2 viral signal, Figures 3D to 3F, throughout the 60-day experimental period is shown below. Sanger sequencing was conducted at time points on day 21, day 45, and day 60 to ensure that the analyses did not produce false positives, with all tests exhibiting a homology greater than 95% to the SARS-CoV-2 reference sequence (Severe acute respiratory syndrome coronavirus 2 genome assembly, chromosome: 1, GenBank: OV387455.1) A statistically significant, unexpected, increase in the measurements of SARS-CoV-2 (121% ± 21%; Figure 3A), PMMoV (75% ± 14%; Figure 3B) and total RNA (248% ± 6%; Figure 3C) at all temperatures are observed at the very beginning of the experiment (between day 0 and day 1 as shown between the grey colour data point at time zero and the subsequent data points shown in blue (T=4°C), orange (T=12°C) and red (T=20°C)). Specifically, increases in SARS-CoV-2 (15% ± 4%), PMMoV (63% ± 10%) and total RNA (160% ± 12%) were recorded across all temperatures. Similarly, an increase at all temperatures for the volume-normalized dataset (25% ± 10%) were also observed while no significant changes were noted for the PMMoV or RNA normalized signals. This increase was also seen in the non-stool-spiked wastewater control, ruling out the possibility this increase was caused solely by the use of spiked-in stool material in this study (Supplementary Figure S2). As further exploration of this increase was beyond the scope of this study, we herein limit the discussion of this phenomenon to a few brief statements that this change may be caused by the stool and wastewater being exposed to the specific simulated bed and near-bed transport conditions in this study, as this same increase was not observed when the stool or the wastewater was exposed to the dynamic suspended transport conditions (i.e. were well mixed throughout the experimental phase). As such, it is possible that the microenvironments within the wastewater mixed with stool created by non-mixed conditions of the vessels throughout the experimental phase may have become anaerobic or anoxic and hence have increased the accessibility to the measurement targets within the wastewater matrix. Indeed, as shown in Figure 4A, a drop of 1.52 ± 0.27 in pH is observed between time 0 and day 1 under bed and near-bed transport conditions while Figure 4B shows that pH remains relatively constant after 1 day of exposure to suspended transport conditions. The measured decrease in pH under simulated bed and near-bed transport conditions supports the potential of anaerobic conditions and a related shift in microenvironments within the wastewater matrix which could in turn lead to a decrease in pH and a change in the partitioning of the target material that may result in an increase of the accessibility of targets during the concentration and extraction analytical processes used in this study (Espinosa et al., 2022). Further work is needed to continue to investigate this unique behaviour of an increase in target material during exposure to unmixed transport conditions.

**Figure 3:**
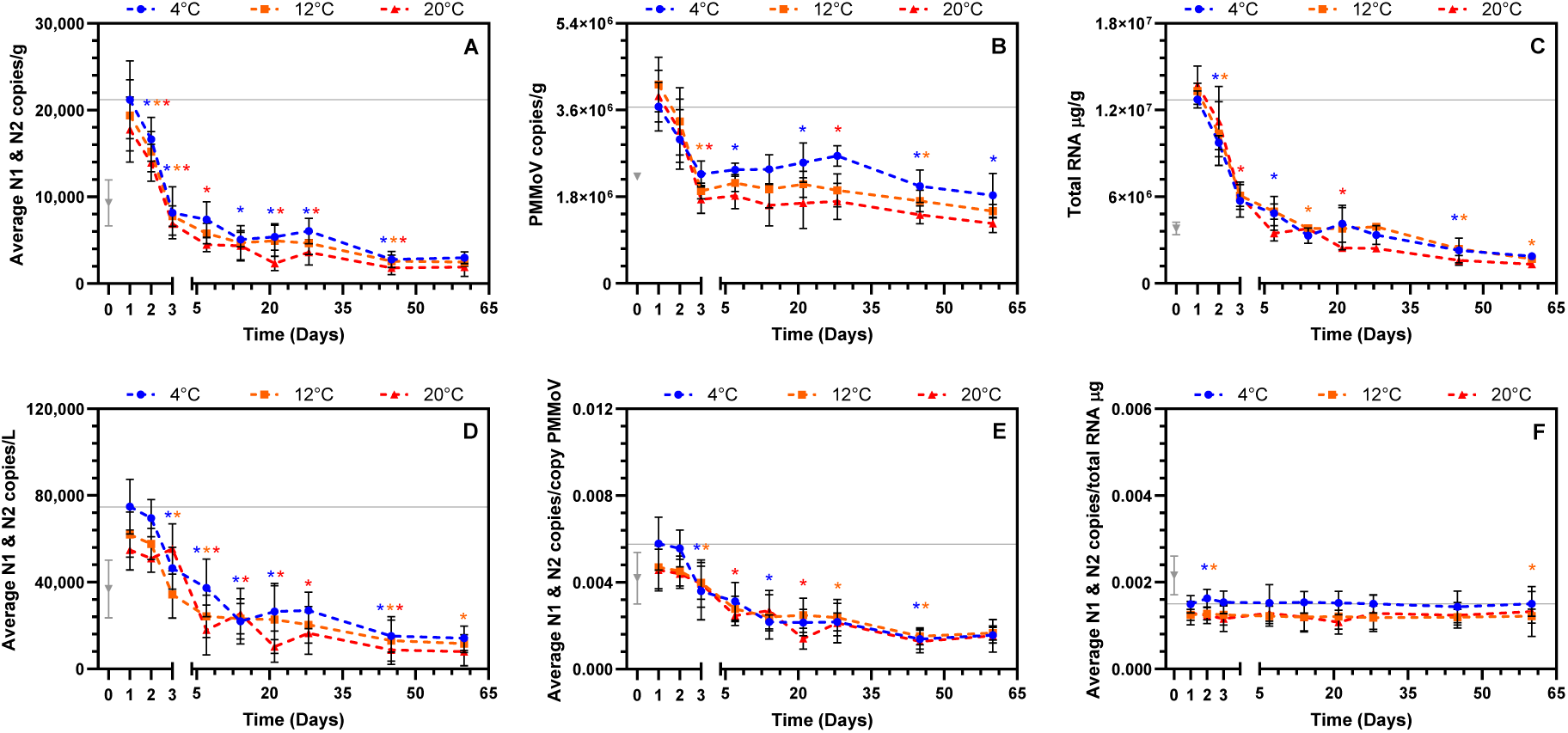
Observed measurements across 60 days under conditions simulating bed and near-bed transport conditions of **A** SARS-CoV-2; **B** PMMoV; **C** total RNA signal. Observed measurements of SARS-CoV-2 across 60 days under conditions simulating bed and near-bed transport conditions normalized by: **D** volume; **E** PMMoV; **F** total RNA. Mean and standard deviations are displayed for three temperatures, 4°C, 12°C and 20°C. Where the standard deviation is too small, the error bars are not displayed. Each measurement is performed in 6 technical triplicates from five biological replicates (n=5). Asterisks indicate which data points are statistically different from the previous one based on p-value cut-off of minimum <0.05.

Decay rates were investigated from day 1 to day 60 of the simulated bed and near-bed transport conditions, following the exclusion of the initial 24-hour increase in signal, which was beyond the scope of this study (Figure 3). Statistically significant decay was observed in SARS-CoV-2 and total RNA signal at all temperatures at day 2 while PMMoV measurements only began showing signs of statistically significant decay at day 3. The heightened stability of PMMoV may be attributed to its robust rod-shaped structure (Kitajima et al., 2014), while SARS-CoV-2 is hypothesized to be present in wastewater primarily as fragmented virions (Kantor et al., 2021). For volume-normalized and PMMoV-normalized signals, statistically significant change was observed on day 3 at 4°C and 12°C, but not 20°C. Total RNA-normalized signal shows no significant changes except on day 2 at 4°C, 12°C and 20°C, due to the unexpected increase described before, and again only on day 60 at 12°C. SARS-CoV-2, PMMoV and total RNA signals as well as volume-normalized signals demonstrated a rapid decay in measured signal between day 1 to 3, followed by a marked tapering of the decay for the remainder of the study (days 7 to 60). During this experiment, PMMoV-normalized viral signal showed a constant decrease between days 1 to 45, while total RNA-normalized viral signal showed almost no change from days 1 to 60. A first order decay model was first investigated to model the experimental data. The mean first order decay rate constants at temperature of 4°C to 20°C for SARS-CoV-2, PMMoV and total RNA ranged from of 0.045 to 0.053 day^−1^, 0.014 to 0.025 day^−1^ and 0.040 to 0.050 day^−1^ respectively. The subsequent calculated decay rates at temperature of 4°C to 20°C for the volume-normalized and PMMoV-normalized signal ranged from, 0.037 to 0.042 day^−1^ and 0.032 to 0.027 day^−1^ respectively. The mean first order decay rate of total RNA-normalized signal was non-calculatable because there as no decay present of this normalized signal.

**Figure 4:**
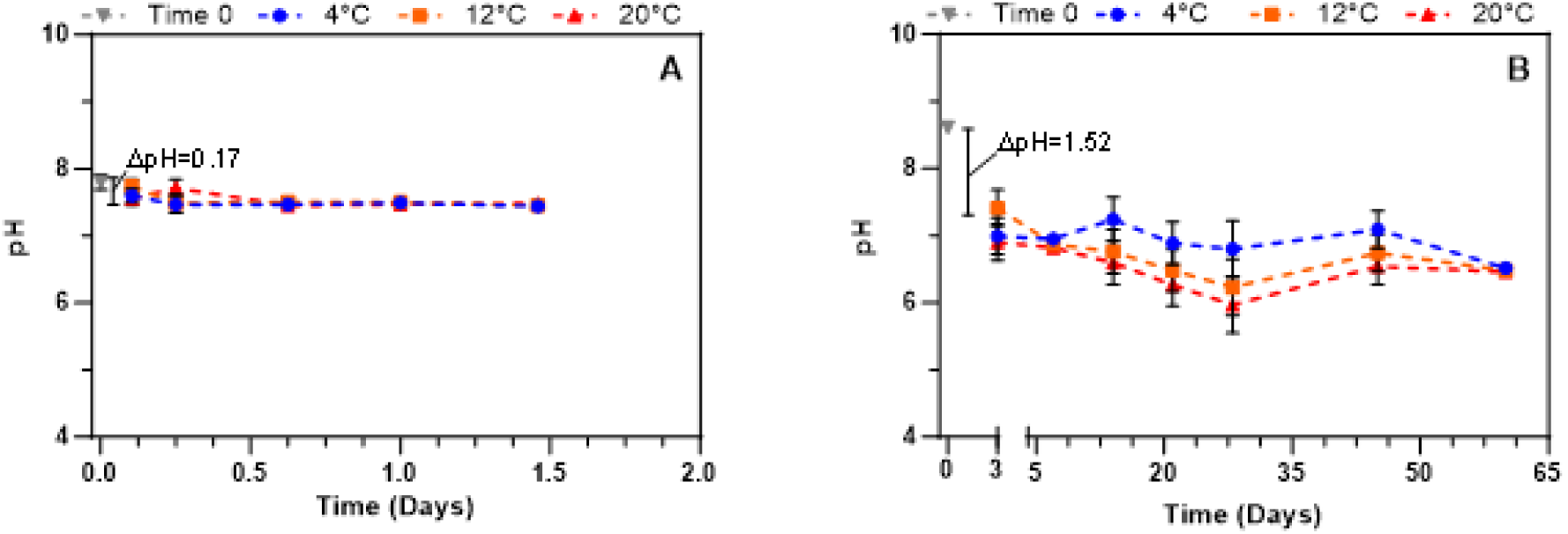
pH variation across time in: **A** conditions simulating dynamic suspended transport; **B** conditions simulating bed and near-bed transport conditions.

The T_90_ values ranged from 43.4 to 51.2 days for SARS-CoV-2, 92.1 to 164.5 days for PMMoV and 46.1 to 57.6 days for total RNA (Table 3). At all temperatures, the SARS-CoV-2 T_90_ observed in this study is larger than the values reported by studies investigating decay rates of lab propagated spiked SARS-CoV-2 material in wastewater (1.6 – 36 days) (Ahmed et al., 2020b; de Oliveira et al., 2021; Hokajärvi et al., 2021). However the reported range significantly broadens (0.5 – 154.9 days) when considering studies investigated decay with endogenous SARS-CoV-2 (Babler et al., 2023; Hart et al., 2023; Roldan-Hernandez et al., 2022; Weidhaas et al., 2021; Yang et al., 2022), with this range now including the T_90_ values measured in this study. The T_90_ values of this study in combination with the findings of previously reported value suggest that endogenous SARS-CoV-2 is more persistent and hence more resistant to decay then lab propagated spiked material. The PMMoV T_90_ observed in this study falls within the range of reported values from other studies investigating endogenous PMMoV material (25.3 – 237.4 days) (Rachmadi, 2016; Roldan-Hernandez et al., 2022; Sala-Comorera et al., 2021). The variability in the reported T_90_ values in current literature suggests that there are additional, unreported factors influencing decay rates. Our study highlights the knowledge gap of the influence of decay from the point of entry into the sewer system and also the influence of various sewer flow conditions on endogenous signal decay. This further emphasizes the need for more research to identify other potential sources of variation, such as wastewater matrix composition, target titer and sewer infrastructure.

**Table 3:**
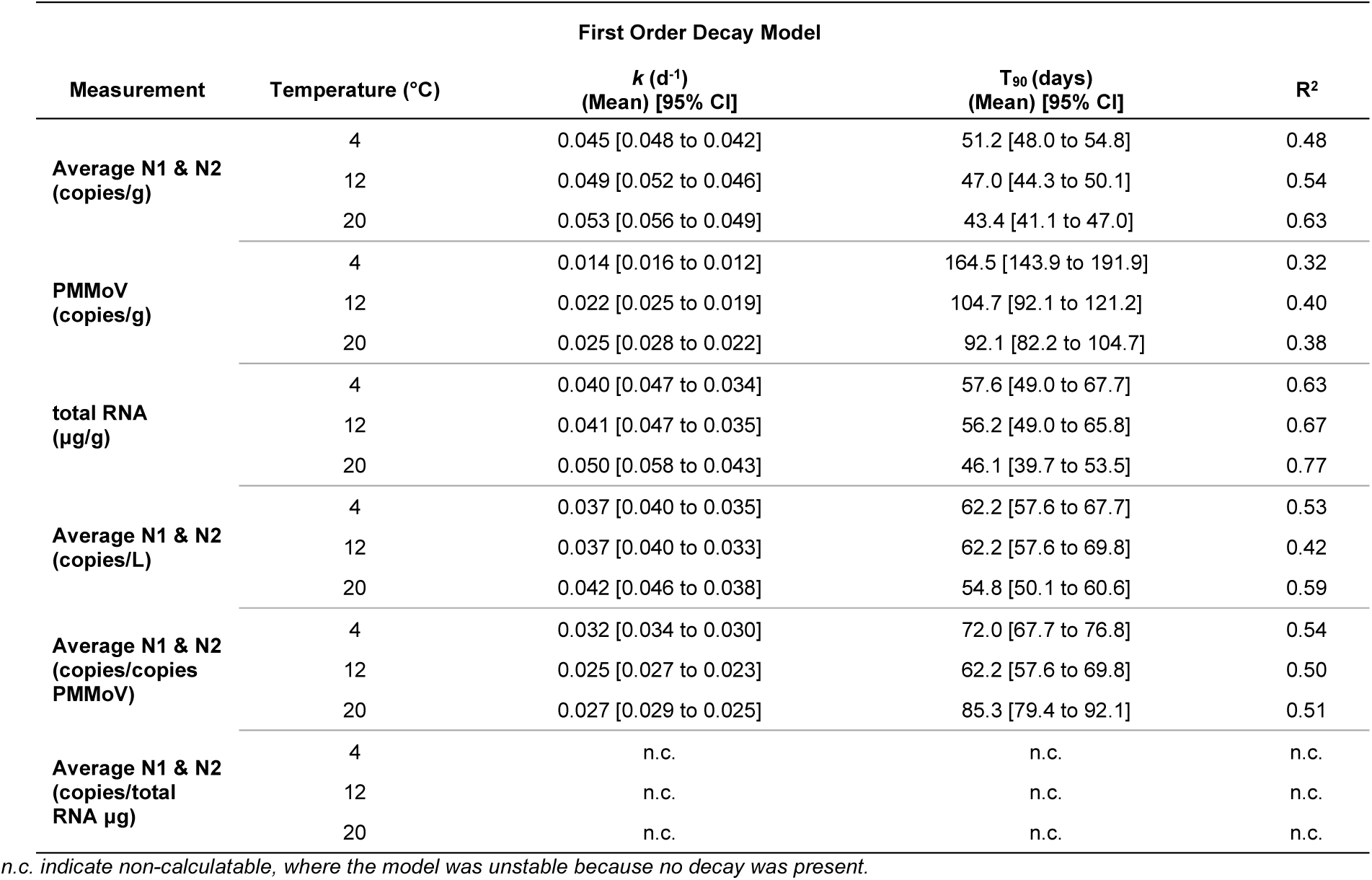
Mean and 95% confidence interval of first order decay constant (k) and T_90_ under conditions simulating toilet flushed stool and bed and near-bed sewer transport.

The three normalization strategies applied in this research, volume-normalized, PMMoV-normalized and total RNA-normalized SARS-CoV-2, demonstrate that the total RNA normalization strategy effectively corrects for time decay of SARS-CoV-2 under bed and near-bed transport conditions, when decay of the viral measurement is significant. An evaluation of whether decay constants were significantly non-zero showed in this study that only the total RNA normalization yielded nonsignificant first order decay constants, with p-values of 0.9588, 0.0511, and 0.1640 at 4°C, 12°C, and 20°C, respectively (Supplemental Table S4). This is to be expected as the decay rate of SARS-CoV-2 was distinct from the decay rate of PMMoV (p<0.001 at all temperatures) while, when compared with total RNA, no significant difference was seen between the decay rate of SARS-CoV-2 and total RNA (p-value of 0.2166, 0.0738 and 0.6824 at 4°C, 12°C and 20°C respectively) (Supplementary Table S5). This suggests that the measurement of SARS-CoV-2 decays at a similar rate to the measurement of the total RNA of stool and wastewaters. Hence, total RNA is identified in this study as an important normalizing marker for sewer decay during bed and near-bed transport conditions and hence also as a potential important indicator of sewer flushing events that are known to re-suspend settled solids from within sewer infrastructure.

#### 3.2.1. Temperature effect

To investigate the effect of temperature on decay rate constants, comparisons were made between the rates at different temperatures: 4°C versus 12°C, 4°C versus 20°C, and 12°C versus 20°C. These comparisons were conducted for measurements of SARS-CoV-2, PMMoV, total RNA, and their normalized values (Supplemental Table S6). A significant temperature effect was observed between the decay rate of SARS-CoV-2 at 4°C and 20°C (p-value=0.0021). The decay rate of PMMoV differed significantly between 4°C and 12°C and between 4°C and 20°C with both showing p-values of less than 0.0001. A significant difference was only observed for the decay rate of total RNA between 4°C and 20°C, with a p-value of 0.0433, which is close to the conventional threshold for significance. This suggests that additional data would be required to more accurately interpret the influence of temperature on total RNA decay. The volume-normalized signal datasets also indicated significant temperature effects, with significant differences observed between 4°C and 20°C (p-value=0.0330), as well as between 12°C and 20°C (p-value=0.0387). Similarly, PMMoV-normalized measurements showed significant differences between 4°C and 12°C, and 4°C and 20°C (p-values<0.0001 and 0.0016, respectively), which could be partially driven by the temperature effect on the normalizer, PMMoV itself. Finally, no temperature effect was observed on the total RNA-normalized signal, as there was no decay detected, which aligns with the finding that total RNA would be a suitable normalizer for sewer decay under bed and near-bed transport conditions and as an identifier of sewer flushing events that re-suspends settled solids. Despite the statistical significance observed in temperature-related differences in decay rates, the actual magnitude of these changes is minimal. When we assess the impact of temperature on the decay constant by linearizing the relationship through a Log_10_ transformation of the mean first order decay rate against temperature, as shown in Figure 5, only PMMoV demonstrates a discernible trend of increasing decay constant with rising temperature. In contrast, SARS-CoV-2, total RNA and volume-normalized Log_10_ linearize decay versus temperature display a flat trend, and the PMMoV-normalized data even shows a slight inverse correlation. This deviates from what is typically reported in the literature for spiked viruses (Ahmed et al., 2020b) where temperature impact is significant. However, this could be attributed to the enhanced persistence of RNA when bound with dissolved organic matter in wastewater (Roldan-Hernandez et al., 2022), which may impede its biodegradation in natural systems, potentially offering protection against temperature effects (Chatterjee et al., 2023). This suggests that the persistent measurement of endogenous SARS-CoV-2, PMMoV, and total RNA is primarily driven by transport conditions and travel time within the sewer system, rather than by temperature. This conclusion is reinforced by the alignment of this study’s decay rates with findings from modelling of endogenous signals, which suggests that time spent in the sewer system has a greater impact on degradation than temperature (Guo et al., 2023; McCall et al., 2022).

**Figure 5:**
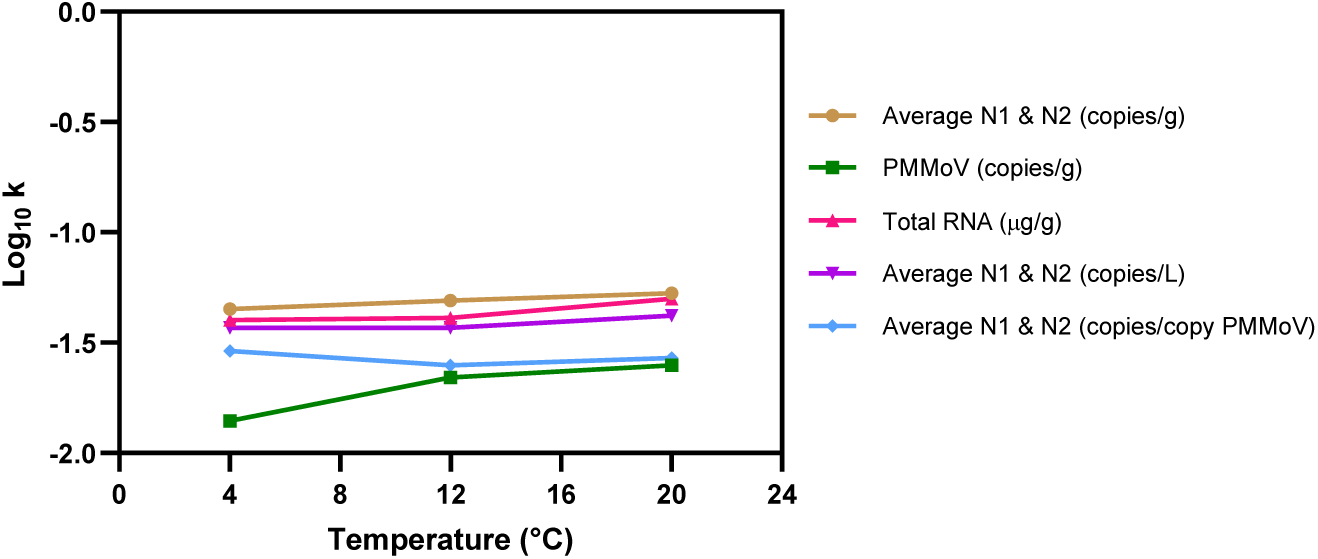
Log10 linearization of the mean first order decay rate constant against temperature.

#### 3.2.1. Two-phase decay

In addition to applying a first order decay model to the experimental data (excluding the initial 24-hour period), a two-phase decay model was also evaluated to describe the decay dynamics observed from day 1 to day 60. The fit of the model was assessed using an extra sum-of-squares F test. For SARS-CoV-2, PMMoV, total RNA signals and the normalized signals, results showed that the F test values exceeded the critical threshold, indicating that the two-phases decay model had a better fit compared to the first order model. This finding implies that decay time might be underestimated by the first order model, with the decrease in measurements being more accurately expressed in the two-phases decay model. The p-values for each test were less than 0.0001, suggesting that the F-statistic results are not likely due to random variability. The total RNA-normalized signal had a p-value of 0.0018, which is significantly higher than other results and because no decay was observable in the RNA-normalized data.

The mean second order decay rates at temperatures ranging from 4°C to 20°C for SARS-CoV-2, PMMoV, and total RNA ranged respectively from 0.660 to 0.842 day^−1^, 0.798 to 1.343 day^−1^, and 0.539 to 0.934 day^−1^ for k_fast_. For k_slow_, these rates were 0.014 to 0.015 day^−1^, 0.004 to 0.005 day^−1^, and 0.016 to 0.018 day^−1^ (Table 4). The calculated decay rates at the same temperature range for the volume-normalized and PMMoV-normalized signals ranged respectively from 0.260 to 0.590 day^−1^ and 0.220 to 0.475 day^−1^ for k_fast_. For k_slow_, these rates were 0.013 to 0.015 day^−1^ and 0.008 to 0.013 day^−1^ (Table 4). The model was unstable and could not fit the RNA-normalized signals as no decay was present. The T_90_ values ranged from 307.2 to 340.7 days for SARS-CoV-2, 1079.9 to 1388.8 days for PMMoV, and 282.8 to 372.6 days for total RNA (Table 4). These values fall outside the range observed in studies using endogenous SARS-CoV-2 (0.5 – 154.9 days) and PMMoV (25.3 – 237.4 days) which could be due to an underestimation of the reported decay rate when modelled with a first order decay model (Roldan-Hernandez et al., 2022; Weidhaas et al., 2021; Yang et al., 2022). This discrepancy may also stem from our study’s approach to measuring persistence and degradation, including the examination of decay from the point of entry into the system, a factor often overlooked in other studies and potentially leading to an underestimation of T_90_ values, and also our approach to simulate common transport conditions within sewer systems. Hence, the use of spiked-in stool samples and assessing the impact of common flow conditions on endogenous SARS-CoV-2 and PMMoV decay could contribute to these observed differences, bridging key knowledge gaps in achieving a more accurate representation of the decay dynamics of these target materials in real-world sewer systems. Once again, the k_fast_ and k_slow_ of SARS-CoV-2 and PMMoV were statistically different, with PMMoV decaying much slower than the SARS-CoV-2 target. On the other hand, the k_fast_ and k_slow_ of total RNA was similar to that of SARS-CoV-2. As a result, both flow and PMMoV were shown not to be adequate normalizers of targets exposed to bed and near-bed transport conditions. These findings suggest that while PMMoV is a good fecal marker normalizer, its slower decay rate makes it unsuitable for normalizing decay occurring in bed and near-bed transport. In addition the study shows that total RNA-normalized signal is an appropriate biomarker to normalize for bed and near-bed transport and in turn is a potential important indicator of sewer flushing events.

**Table 4:** Mean and 95% confidence interval of two-phase decay constant (k) and T_90_ under conditions simulating toilet flushed stool and bed and near-bed sewer transport.

| Two-Phase Decay Model |  |  |  |  |  |  |
| --- | --- | --- | --- | --- | --- | --- |
| Measurement | Temperature (°C) | $k$ (day<sup>-1</sup>)<br>(Mean) [95% CI] | $T_{90}$ (days) [95% CI] | Fast phase<br>Percentage (%) | R<sup>2</sup> | |
| Average N1 & N2<br>(copies/g) | 4 | $k_{fast}$ : 0.842 [0.712 to 1.000] | 340.7 [464.6 to 255.6] | 81.7 [78.8 to 84.2] | 0.79 | |
| | | $k_{slow}$ : 0.015 [0.011 to 0.020] | | | | |
| | 12 | $k_{fast}$ : 0.700 [0.603 to 0.812] | 363 [564.6 to 267.4] | 82.4 [79.9 to 84.7] | 0.84 | |
| | | $k_{slow}$ : 0.014 [0.009 to 0.019] | | | | |
| | 20 | $k_{fast}$ : 0.660 [0.576 to 0.756] | 307.2 [491.6 to 223.4] | 85.7 [83.3 to 87.8] | 0.86 | |
| | | $k_{slow}$ : 0.016 [0.010 to 0.022] | | | | |
| PMMoV (copies/g) | 4 | $k_{fast}$ : 1.343 [0.751 to 3.594] | 1388.8 [2777.5 to 793.6] | 63.2 [47.2 to 90.7] | 0.54 | |
| | | $k_{slow}$ : 0.004 [0.002 to 0.007] | | | | |
| | 12 | $k_{fast}$ : 0.875 [0.634 to 1.213] | 1088.2 [2720.6 to 680.1] | 70.6 [63.6 to 76.7] | 0.76 | |
| | | $k_{slow}$ : 0.005 [0.002 to 0.008] | | | | |
| | 20 | $k_{fast}$ : 0.789 [0.556 to 1.108] | 1079.9 [2699.7 to 599.9] | 72.6 [65.6 to 78.4] | 0.74 | |
| | | $k_{slow}$ : 0.005 [0.002 to 0.009] | | | | |
| total RNA<br>(µg/g) | 4 | $k_{fast}$ : 0.934 [0.707 to 1.269] | 327.6 [476.5 to 238.3] | 78.3 [73.6 to 82.4] | 0.92 | |
| | | $k_{slow}$ : 0.016 [0.011 to 0.022] | | | | |
| | 12 | $k_{fast}$ : 0.922 [0.536 to 1.831] | 299.8 [539.6 to 199.9] | 72.7 [62.1 to 84.1] | 0.89 | |
| | | $k_{slow}$ : 0.018 [0.010 to 0.027] | | | | |
| | 20 | $k_{fast}$ : 0.539 [0.389 to 0.746] | 282.8 [1272.8 to 149.7] | 82.2 [74.6 to 88.1] | 0.63 | |
| | | $k_{slow}$ : 0.018 [0.004 to 0.034] | | | | |
| Average N1 & N2<br>(copies/L) | 4 | $k_{fast}$ : 0.449 [0.317 to 0.626] | 394.2 [613.2 to 275.9] | 66.1 [59.6 to 71.4] | 0.68 | |
| | | $k_{slow}$ : 0.014 [0.009 to 0.020] | | | | |
| | 12 | $k_{fast}$ : 0.590 [0.445 to 0.775] | 416.3 [676.5 to 284.8] | 71.9 [65.7 to 77.0] | 0.61 | |
| | | $k_{slow}$ : 0.013 [0.008 to 0.019] | | | | |
| | 20 | $k_{fast}$ : 0.260 [0.189 to 0.350] | 357.4 [1340.2 to 206.2] | 74.2 [64.8 to 82.2] | 0.68 | |
| | | $k_{slow}$ : 0.015 [0.004 to 0.026] | | | | |
| Average N1 & N2<br>(copies/copies<br>PMMoV) | 4 | $k_{fast}$ : 0.475 [0.325 to 0.667] | 427.7 [695 to 308.9] | 62.7 [56.5 to 67.9] | 0.69 | |
| | | $k_{slow}$ : 0.013 [0.008 to 0.018] | | | | |
| | 12 | $k_{fast}$ : 0.319 [0.217 to 0.463] | 562.7 [937.8 to 401.9] | 50.1 [42.2 to 56.8] | 0.61 | |
| | | $k_{slow}$ : 0.010 [0.006 to 0.014] | | | | |
| | 20 | $k_{fast}$ : 0.220 [0.138 to 0.334] | 697.9 [5582.9 to 398.8] | 60.4 [51.3 to 69.3] | 0.63 | |
| | | $k_{slow}$ : 0.008 [0.001 to 0.014] | | | | |
| Average N1 & N2<br>(copies/total RNA<br>µg) | 4 | $k_{fast}$ : n.c. | n.c. | n.c. | n.c. | |
| | | $k_{slow}$ : n.c. | | | | |
| | 12 | $k_{fast}$ : n.c. | n.c. | n.c. | n.c. | |
| | | $k_{slow}$ : n.c. | | | | |
| | 20 | $k_{fast}$ : n.c. | n.c. | n.c. | n.c. | |
| | | $k_{slow}$ : n.c. | | | | |

n.c. indicate non-calculatable, where the model was unstable because no decay was present.

## 4. Conclusions

Our study offers insights into the endogenous decay of SARS-CoV-2, PMMoV and total RNA from point of entry in the sewer system, the toilet flush event, and during two predominant sewer transport conditions. Simulated dynamic suspended transport over 35 hours period revealed good persistence and minimal degradation of the measurement of SARS-CoV-2, PMMoV, and total RNA throughout short, moderate and long sewer transport conditions that simulate small, medium and large sewersheds subsections. The observed decay rates showed no significant decay rate for SARS-CoV-2, PMMoV or total RNA which appears to be independent of temperature effects within the temperature range of 4°C to 20°C. This finding indicates negligible decay in dynamic suspended transport.

In contrast, the experiments simulating bed and near-bed transport conditions for 60 days demonstrated an initial unexpected increase in the measurement of SARS-CoV-2, PMMoV, and total RNA. This could be attributed to microenvironment shifts caused by the simulated transport conditions, a phenomenon that warrants further investigation. Due to the complexity of this initial phase, the data from day 0 to 1 were excluded from the decay analysis. Subsequently, decay was computed from day 1 to 60, where significant decay rates were observed with differing decay patterns for SARS-CoV-2, PMMoV, and total RNA being observed. Temperature effect was minimal, suggesting the decay is primarily driven by transport conditions and travel time within the sewer system, rather than by temperature. The decay rates of the simulated bed and near-bed transport were observed to be within the range of previous studies on endogenous targets. Although within the range, the variability in reported decay patterns suggests the potential influence of other parameters like wastewater matrix composition, viral titers, or sewer system dynamics, which need further exploration. To that end, our research particularly highlighted the previously overlooked impacts on endogenous signals of decay from point of entry into the system and the role of different flow conditions in this process. While our first order decay model fell short in predicting the decay rates, a two-phases decay model significantly improved the fit during bed and near-bed transport. Total RNA normalization emerged as the most effective strategy for correcting time decay in sewer systems experiencing bed and near-bed transport conditions. The outcomes of our study have implications for understanding and modelling of SARS-CoV-2 WBS in sewersheds, especially systems that undergo bed and near-bed transport conditions followed by sudden resuspension and mixing events, such as large rainfall events, where flushing of the sewer infrastructure causes the re-suspension of SARS-CoV-2, PMMoV and total RNA gene targets. This study also underlines the need for further investigation into time zero, toilet flush decay studies performed under different sewer transport conditions.

## Data Availability

All data produced in the present study are available upon reasonable request to the authors

## 6. Acknowledgements

The authors wish to acknowledge the help and assistance of the University of Ottawa, the Ottawa Hospital, the Children’s Hospital of Eastern Ontario, the Children’s Hospital of Eastern Ontario’s Research Institute, Public Health Ontario and all their employees involved in the project. Most specifically, the authors wish to thank Jessica Haines, Rebecca Porteous, Irene Watpool. Their time, facilities, resources, and feedback are greatly appreciated.

